# HIV and antiretroviral treatment as drivers of rifampicin-resistant tuberculosis in South Africa: insights from mathematical modelling

**DOI:** 10.64898/2025.12.11.25342033

**Authors:** Leigh F. Johnson, Lukas Fenner, Pren Naidoo, Mmamapudi Kubjane, Denise Evans, Shaheed V. Omar, Nesbert Zinyakatira, Helen Cox

## Abstract

**Background:** High levels of multidrug and rifampicin resistant tuberculosis (MDR/RR-TB) are a global concern, although they have declined over the last decade. TB patients are at increased risk of acquired rifampicin resistance if they have HIV coinfection, especially at low CD4 counts, but this dynamic has not previously been modelled.

**Methods:** We extended a previously-developed model that simulates HIV and TB in South African adults, to include the acquisition and transmission of rifampicin resistance. In line with systematic reviews, the risk of acquiring RR with TB treatment is modelled as being negatively associated with patients’ CD4 counts. We allow for temporal changes in drug susceptibility testing, both before treatment initiation and at treatment failure, as well as other changes in TB prevention and treatment. The model is calibrated to data from national TB drug-resistance surveys, and recorded numbers of MDR/RR-TB laboratory diagnoses and patients initiating second-line TB treatment, using a Bayesian approach.

**Results:** The model estimates that the proportion of South African TB patients with rifampicin resistance at diagnosis increased from 2.0% (95% CI: 1.7-2.3%) in 1986 to 5.9% (5.2-6.9%) in 2013, in line with survey data. In the absence of HIV, the prevalence of MDR/RR-TB would have increased to 4.1% (2.7-5.1%) in 2013, suggesting a third of rifampicin resistance in 2013 was attributable to HIV. In the absence of antiretroviral treatment (ART), the prevalence of rifampicin resistance would have been higher (6.5% [5.6-7.6%] in 2013, rising to 6.9% [5.7-8.2%] in 2019). ART reduced the prevalence of rifampicin resistance in 2019 by 17%.

**Conclusions:** In countries with high HIV prevalence, HIV may be a major driver of rifampicin resistance in people with TB. ART programmes have the potential to reduce the emergence of resistance substantially.

## Background

Drug-resistant tuberculosis (TB) is a major public health concern. TB that is resistant to rifampicin and isoniazid, the two most commonly used first-line drugs, is strongly associated with increased risk of mortality [1, 2] and treatment failure [3]. In addition, drug-resistant TB is substantially more expensive to treat [4], and typically requires longer durations of treatment. Multidrug-resistant TB (MDR-TB, which is resistant to both rifampicin and isoniazid) is very frequent in people with rifampicin-resistant TB [1, 5], and treatment guidelines therefore place particular emphasis on testing for rifampicin resistance.

Globally, the incidence of rifampicin-resistant TB (RR-TB) has declined over the last decade, both in absolute terms and as a fraction of incident TB [6], despite early modelling studies predicting a steady rise in the fraction of TB cases that are RR-TB [7, 8]. This is likely to be a reflection of major progress in scaling up RR-TB screening and treatment [6]. Nevertheless, approximately 390 000 people developed RR-TB in 2024, 4% of all people developing TB [6].

South Africa has one of the highest TB incidence rates in the world, with the adult TB incidence rate estimated at 641 per 100 000 adults over 2023-24 [9]. The TB epidemic over the last three decades has been exacerbated by high levels of HIV prevalence, although this has been mitigated partially by the introduction of antiretroviral treatment (ART) [10]. National surveys estimate that the proportion of TB that was RR-TB increased from 1.8% in the 1980s [11] to 3.4% in 2001-02 [12] and 4.6% in 2012-14 [5]. This represents a major challenge, not least because the costs of managing drug resistance make up two thirds of public spending on TB in South Africa [4]. The explanations for the rise in RR-TB are unclear, and there is also some uncertainty regarding trends in the last decade. Some increases in resistance may have been due to increased levels of screening and treatment for TB [10]. It is also possible that some of the increase in RR-TB, as a proportion of all TB, has been due to rising levels of HIV-related immune suppression, which is a major risk factor for the acquisition of rifampicin resistance while on treatment [13-17].

In this study we aim to assess the explanations for the rising RR-TB prevalence in South Africa, and to evaluate the trends in the period after the most recent survey in 2012-14. We address these objectives by extending a previously developed model of TB and HIV in South Africa [10] and calibrating the model to a combination of survey data and routine TB notification and testing data.

## Methods

### Model structure

We extended the Thembisa TB model to include RR-TB. The previous version of the model, which did not differentiate between drug-susceptible and drug-resistant TB, is described elsewhere [10, 18], and is represented by the first two columns in Figure 1. Briefly, individuals acquire *Mycobacterium tuberculosis* (MTB) infection at a rate that depends on their age- and sex-specific rate of social contact and the prevalence of active TB in their social contacts. A proportion of those who acquire MTB develop active TB immediately, while the rest naturally clear the infection or remain latently infected; some of the latter may develop active TB years later (‘reactivation’). People with active TB are classified as smear-negative or smear-positive, with the latter having higher infectiousness and more frequent symptoms. Symptomatic individuals seek treatment and are microbiologically tested at a rate determined by published numbers of tests and diagnoses [19]. A proportion of those diagnosed positive initiate treatment, and symptomatic TB patients can also be treated empirically in the absence of microbiological diagnosis. Standard treatment is assumed to last for 6 months, and of those who complete treatment, a high proportion are assumed to be cured; in those who discontinue treatment before 6 months, a lower proportion is cured. Those who are not cured are assumed to return to the untreated active TB state. In those who are cured, there is assumed to be a high risk of relapse during the first 6 months after treatment completion. People who are HIV-positive can initiate TB preventive therapy (TPT), which is assumed to reduce active TB incidence. The population is further stratified by age, sex and HIV status/stage, with HIV-positive adults being classified according to their CD4 stage (500+, 350-499, 200-349 or <200 cells/*μ*l) and receipt of ART. The model of HIV and TB in South Africa begins in 1985, with numbers in different HIV and TB states being updated at monthly time steps. Only adult TB (ages 15 and older) is represented.

**Figure 1.**
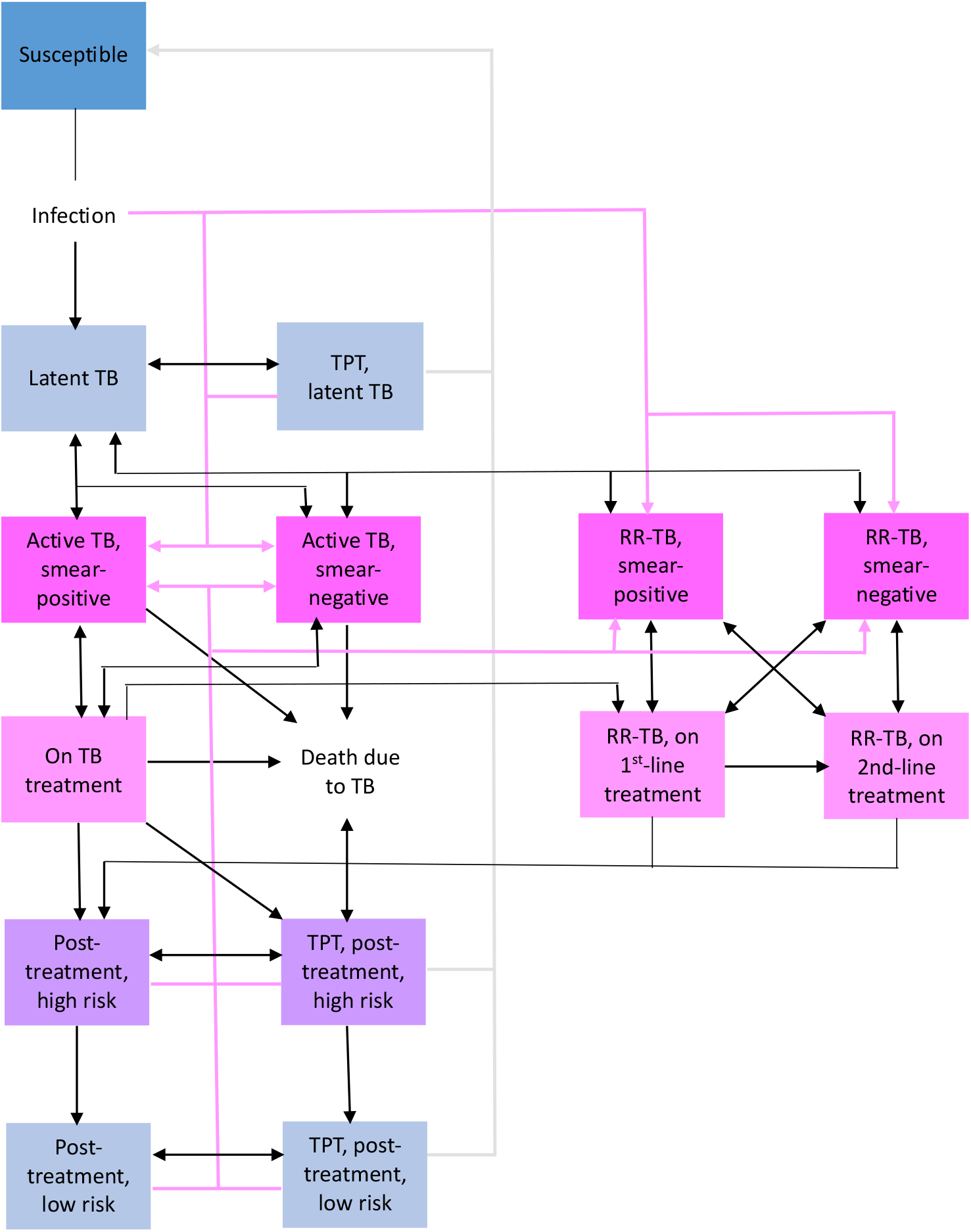
Adult TB model structure Pink lines represent ‘fast progression’ to active TB (soon after *Mycobacterium tuberculosis* infection or reinfection). Solid grey lines represent return to the susceptible state in people who are successfully treated with TB preventive therapy (TPT). RR-TB = rifampicin-resistant TB.

For the purpose of this study, the model structure was extended to include four additional states (represented in the last two columns of Figure 1): two states representing untreated RR-TB (one smear-positive, one smear-negative), and two states representing treated RR-TB (one for patients inappropriately treated with first-line drugs, the other for patients receiving appropriate second-line treatment).

The calculation of total TB incidence is the same as described previously [10]. To represent the transmission of resistant TB, the proportion of incident TB cases that are rifampicin-resistant in year *t* is calculated as

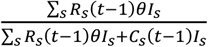

where *R*_*s*_(*t* − 1) and *C*_*s*_(*t* − 1) represent the number of rifampicin-resistant and other TB cases respectively, of smear status *s*, in the previous year, *θ* represents the relative infectivity of RR-TB cases (relative to others) and *I*_*s*_ represents the relative infectivity of TB patients with smear status *s* (relative to smear-positive TB). To represent the acquisition of resistance, we assume that patients who are receiving first-line TB treatment develop rifampicin resistance with probability *π* if treatment fails, consistent with the parameterization in other models [20, 21]. In HIV-positive individuals the probability of developing rifampicin resistance if treatment fails is

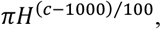

where *H* is the factor by which the probability of acquired resistance reduces, per 100-cell increase in the individual’s CD4 count, and *c* is the HIV-positive individual’s CD4 count (HIV-negative individuals are assumed to have an average CD4 count of around 1000 cells/μl).

### Diagnosis and treatment of RR-TB

Prior to the introduction of GeneXpert testing in 2011, drug susceptibility testing (DST) was conducted using culture or Line Probe Assay. South African guidelines recommended DST only if patients had a history of previous TB treatment, were contacts of MDR-TB patients or were failing to respond to first-line treatment [22]. We calculate the probability that a treated RR-TB case receives DST as

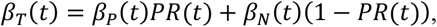

where *β*_*P*_(*t*) is the proportion of previously-treated patients who receive DST in year *t, β*_*N*_(*t*) is the proportion of new patients (not previously treated) who receive DST, and *PR*(*t*) is the proportion of people with RR-TB treated in year *t* who were previously treated. The estimation of *PR*(*t*) is explained in the appendix. Table 1 shows the assumed values of *β*_*P*_(*t*) and *β*_*N*_(*t*), based on South African studies [23-25]. As GeneXpert automatically tests for rifampicin resistance, we assume that DST is conducted in 100% of all patients tested by Xpert, and thus the assumed proportions receiving DST in the period after the Xpert rollout are calculated as a function of the Xpert rollout. In the period 2001-2008, the rates of DST are assumed to be the same as in 2008-09, and in the period before 2001, the DST rates are calculated by multiplying the 2008-09 rates by a factor of *W*_0_⁄*W*_1_ (defined below). These rates of testing are assumed to apply only to patients who are microbiologically diagnosed, i.e. patients who are empirically treated are assumed not to get DST. People with RR-TB who receive DST or GeneXpert are assumed to initiate second-line drugs; the rest initiate first-line treatment.

**Table 1:** Assumed proportions of TB cases that receive drug susceptibility testing.

| Year | 2008-09 | 2009-10 | 2010-11 | 2011-12 | 2012-13 | 2013-14 | 2014-15 | Post-2015 |
| --- | --- | --- | --- | --- | --- | --- | --- | --- |
| Previously treated | 27.2% | 47.6% | 68.0% | 75.0% | 81.8% | 88.8% | 91.4% | 93.6% |
| Newly treated | 6.7% | 19.4% | 32.0% | 47.0% | 61.2% | 76.2% | 81.6% | 86.4% |

DST is also assumed to be conducted in patients who are not responding to first-line TB treatment. We assume that in year *t* a proportion of patients who are not responding to first-line treatment get DST, this proportion being *W*(*t*) in the patients who continue treatment and *kW*(*t*) in patients who discontinue treatment. (In the former group, those who test rifampicin-resistant automatically get switched to second-line treatment, while in the latter group we assume treatment is discontinued completely.) Due to lack of data, we represent the trend in *W*(*t*) using a simple ‘step function’, assuming increases following the 2001 South African treatment guidelines and the 2011 guidelines [26]:

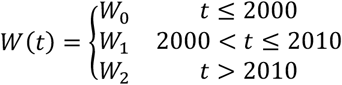

The duration of second-line treatment is assumed to be 18 months in the period to 2018 [27], and 9 months after 2018, following the introduction of bedaquiline [28]. In patients who receive second-line treatment, the monthly rates of mortality and treatment discontinuation are assumed to be the same as for patients with drug-sensitive TB, and probabilities of treatment failure after completing/discontinuing treatment are also the same as for patients with drug-sensitive TB. However, RR-TB patients who are inappropriately treated with first-line drugs are assumed to have a monthly mortality rate 7 times that in drug-sensitive TB patients [1, 2], and their odds of treatment failure are assumed to be higher than those in drug-sensitive TB patients, by a factor of *Ψ*.

### Model calibration

The model is calibrated to three different national data sources: surveys of RR-TB prevalence in 2001-02 [12] and 2012-14 [5]; National Institute for Communicable Diseases (NICD) data on numbers of laboratory specimens with positive RR-TB results over 2010-21 [29]; and numbers of people initiating second-line treatment over 2007-2020 [26, 29]. We adopted a Bayesian approach to model calibration. In the first step, we specified prior distributions to represent the uncertainty in key model parameters (Table 2). In the case of the DST parameters for patients not responding to first-line treatment, we lacked appropriate data, but assumed that rates of DST would have increased over time, and would have been lower in patients discontinuing treatment than in those completing/continuing treatment. We therefore specified vague priors (uniform on the interval [0, 1]) to represent the uncertainty in *W*_2_ and *k*, as well as in the ratios *W*_1_⁄*W*_2_ and *W*_0_⁄*W*_1_. Beta priors were assigned to all other parameters (except *θ* and *Ψ*, for which gamma priors were specified). All other TB and HIV parameters not shown in Table 2 were fixed at the values estimated previously [18].

**Table 2:**
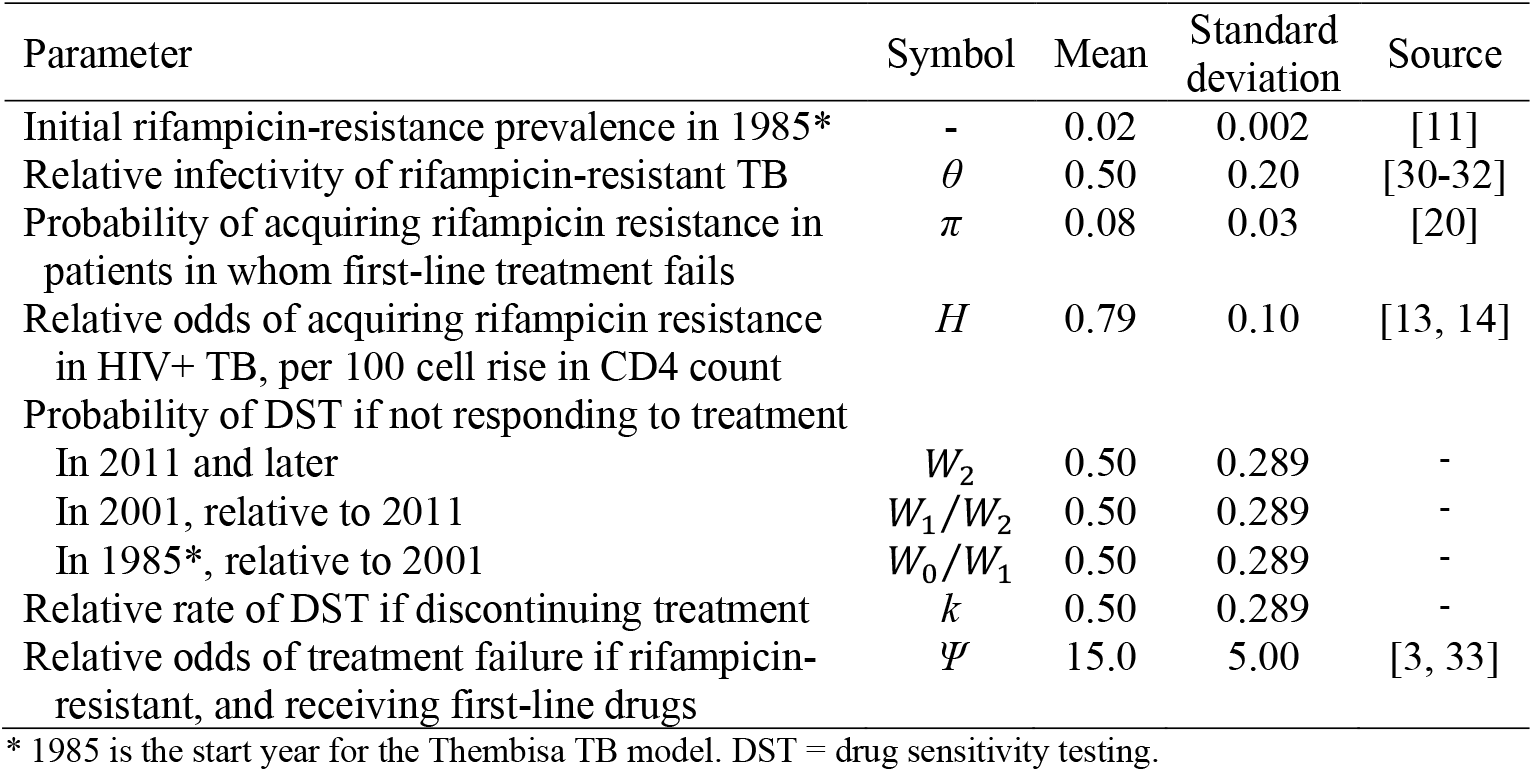
Prior distributions.

| Parameter | Symbol | Mean | Standard deviation | Source |
| --- | --- | --- | --- | --- |
| Initial rifampicin-resistance prevalence in 1985* | - | 0.02 | 0.002 | [11] |
| Relative infectivity of rifampicin-resistant TB | $\theta$ | 0.50 | 0.20 | [30-32] |
| Probability of acquiring rifampicin resistance in patients in whom first-line treatment fails | $\pi$ | 0.08 | 0.03 | [20] |
| Relative odds of acquiring rifampicin resistance in HIV+ TB, per 100 cell rise in CD4 count | $H$ | 0.79 | 0.10 | [13, 14] |
| Probability of DST if not responding to treatment |  |  |  |  |
| In 2011 and later | $W_2$ | 0.50 | 0.289 | - |
| In 2001, relative to 2011 | $W_1/W_2$ | 0.50 | 0.289 | - |
| In 1985*, relative to 2001 | $W_0/W_1$ | 0.50 | 0.289 | - |
| Relative rate of DST if discontinuing treatment | $k$ | 0.50 | 0.289 | - |
| Relative odds of treatment failure if rifampicin-resistant, and receiving first-line drugs | $\Psi$ | 15.0 | 5.00 | [3, 33] |

\* 1985 is the start year for the Thembisa TB model. DST = drug sensitivity testing.

In the second Bayesian step, we specified likelihood functions to represent the extent of consistency between the model outputs and the calibration data, taking into account potential biases in the data. Prevalence survey data were adjusted to account for under-estimation of RR-TB due to reliance on phenotypic detection methods that missed borderline resistant mutations that were not considered defining of resistance at the time of the surveys, but which are now included in the WHO definition of rifampicin resistance. Bias in the NICD data could arise due to double counting of multiple tests in the same individual, or imperfect test sensitivity and specificity. Bias in the numbers of second-line treatment initiations could arise due to under-reporting in the electronic record system. Further details regarding the likelihood definition are provided in the appendix.

In the third Bayesian step, the posterior distribution of parameters most consistent with the prior distributions and likelihood function was estimated numerically, using Incremental Mixture Importance Sampling [34]. A sample of 1000 parameter combinations was drawn from the posterior distribution, and detailed model results were generated for each parameter combination. The results presented are the means from the 1000 parameter combinations (and and 97.5 percentiles where 95% confidence intervals [CIs] are presented).

### Scenarios

To assess the significance of different drivers of drug resistance, we compared the model estimates of past trends in drug resistance based on actual programme data (‘actual’) against a number of counterfactual scenarios. These included (1) a scenario in which there was no HIV epidemic; (2) a scenario in which there was no introduction of ART; (3) a scenario in which microbiological testing rates in people with symptomatic TB remained constant after 2004 instead of increasing; and (4) a scenario in which there was no increase in DST after 2008 in people presenting with TB symptoms. The first three counterfactual scenarios are the same as described in our previous work [10].

## Results

### Comparison of prior and posterior distributions

Table 3 compares the prior and posterior distributions. Posterior estimates of the relative infectivity of rifampicin-resistant TB are higher than the prior estimates (mean of 0.84 compared to 0.5), and closer to the value of 0.72 assumed for MDR-TB in a number of other modelling studies [7, 35]. Other posterior means are reasonably close to the prior means.

**Table 3:** Comparison of prior and posterior distributions.

| Parameter | Prior<br>(mean, 95% CI) | Posterior<br>(mean, 95% CI) |
| --- | --- | --- |
| Initial rifampicin-resistance prevalence | 0.020 (0.016-0.024) | 0.020 (0.017-0.023) |
| Relative infectivity of rifampicin-resistant TB | 0.500 (0.188-0.962) | 0.836 (0.755-0.891) |
| Probability of acquiring rifampicin resistance in patients in whom first-line treatment fails | 0.080 (0.032-0.148) | 0.087 (0.053-0.135) |
| Relative odds of acquiring rifampicin resistance in HIV+ TB, per 100 cell rise in CD4 count | 0.790 (0.564-0.946) | 0.832 (0.716-0.919) |
| Probability of DST if not responding to treatment |  |  |
| In 2011 and later | 0.500 (0.025-0.975) | 0.375 (0.152-0.694) |
| In 2001, relative to 2011 | 0.500 (0.025-0.975) | 0.617 (0.336-0.941) |
| In 1985*, relative to 2001 | 0.500 (0.025-0.975) | 0.540 (0.055-0.921) |
| Relative rate of DST if discontinuing treatment | 0.500 (0.025-0.975) | 0.549 (0.035-0.970) |
| Relative odds of treatment failure if rifampicin-resistant, and receiving first-line drugs | 15.00 (6.86-26.27) | 12.06 (6.34-20.57) |

### Calibration to rifampicin resistance data

Figure 2 compares the model estimates of the proportion of TB cases that are rifampicin-resistant against the survey data that are used in calibrating the model. The proportion of South African TB patients with rifampicin resistance at diagnosis increased from 2.0% (95% CI: 1.7-2.3%) in 1986 to 5.9% (5.2-6.9%) in 2013. The model estimates are in reasonably good agreement with the calibration data, although the model estimates are slightly higher than the survey estimates over the 2012-14 period.

**Figure 2.**
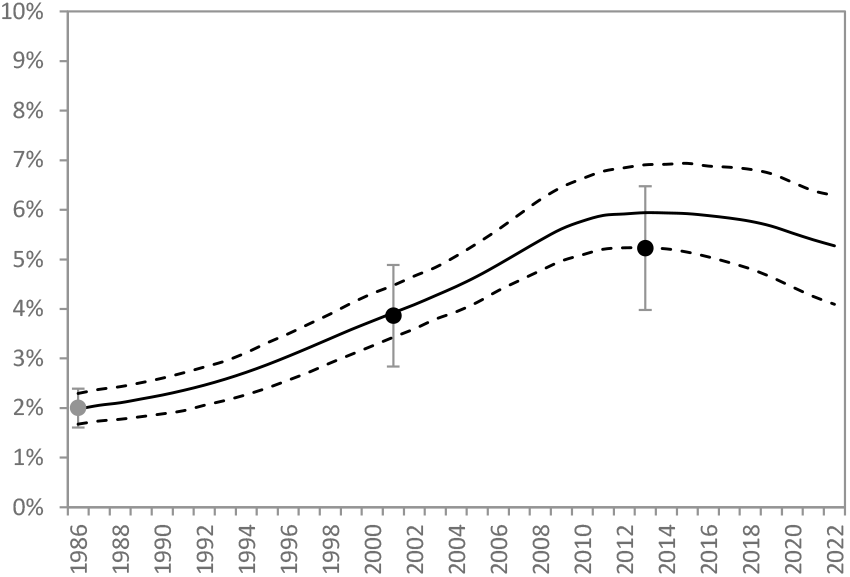
Proportion of adult TB cases who are rifampicin-resistant Survey data (dots) have been adjusted to account for an assumed 88% sensitivity. The 1986 data point was not used in calibration (as it only informed the prior distribution for the initial prevalence of rifampicin resistance), and is therefore formatted in grey. The solid line represents the posterior mean of the model estimates (the proportion of untreated adult TB that is rifampicin-resistant) and the dashed lines represent 95% credible intervals.

Figure 3 compares the model estimates of numbers of new detected cases of RR-TB and numbers of patients starting second-line treatment against routine data sources. The routine data have been adjusted to reflect the estimated levels of bias: in the case of reported numbers of rifampicin-resistant diagnoses, we estimate the reported number to be 31% (95% CI: 10-47%) greater than the true number, suggesting problems with specificity and/or under-matching in the NHLS de-duplication algorithm. However, the reported number of second-line treatment initiations are estimated to be 6% (95% CI: 0-21%) less than the true number, suggesting relatively limited undercount in the recorded numbers starting second-line treatment. Reported levels of rifampicin resistant diagnosis peak more rapidly than our model suggests (in 2012) and also decline more dramatically. Reported numbers of second-line treatment initiations rise later than our model suggests (in 2012-14), but decline in line with our model estimates after 2014.

**Figure 3.**
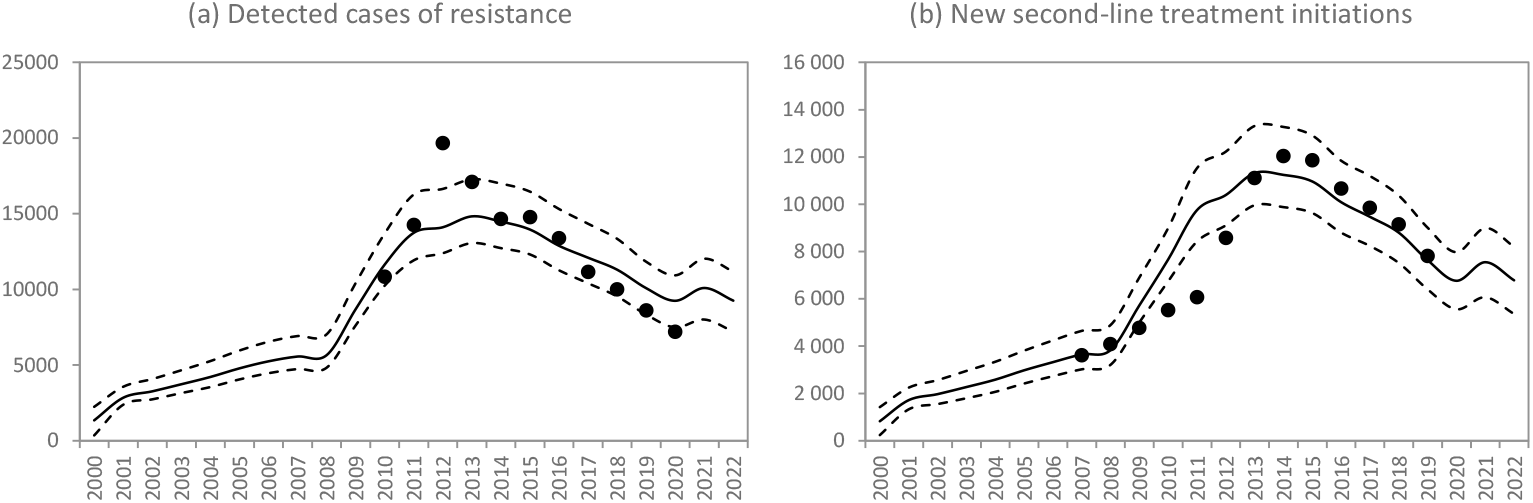
Numbers of diagnosed rifampicin-resistant cases and adults starting second-line TB treatment NICD and WHO estimates (based on routine reporting) are represented by dots, after adjustment for estimated levels of under-/over-reporting. The model estimates are represented by the solid lines (posterior means) and dashed lines (95% prediction intervals).

### Counterfactual scenarios

Figure 4 shows the proportion of prevalent TB that is rifampicin-resistant, under various different scenarios. Figure 4a shows that in the absence of HIV, the prevalence of RR-TB would have increased to 4.1% (2.7-5.1%) in 2013, suggesting a third of rifampicin resistance in 2013 was attributable to HIV. However, in the absence of ART the prevalence of rifampicin resistance would have been higher (6.5% [5.6-7.6%] in 2013, rising to 6.9% [5.7-8.2%] in 2019). ART therefore reduced the prevalence of rifampicin resistance in 2019 by 17%.

**Figure 4.**
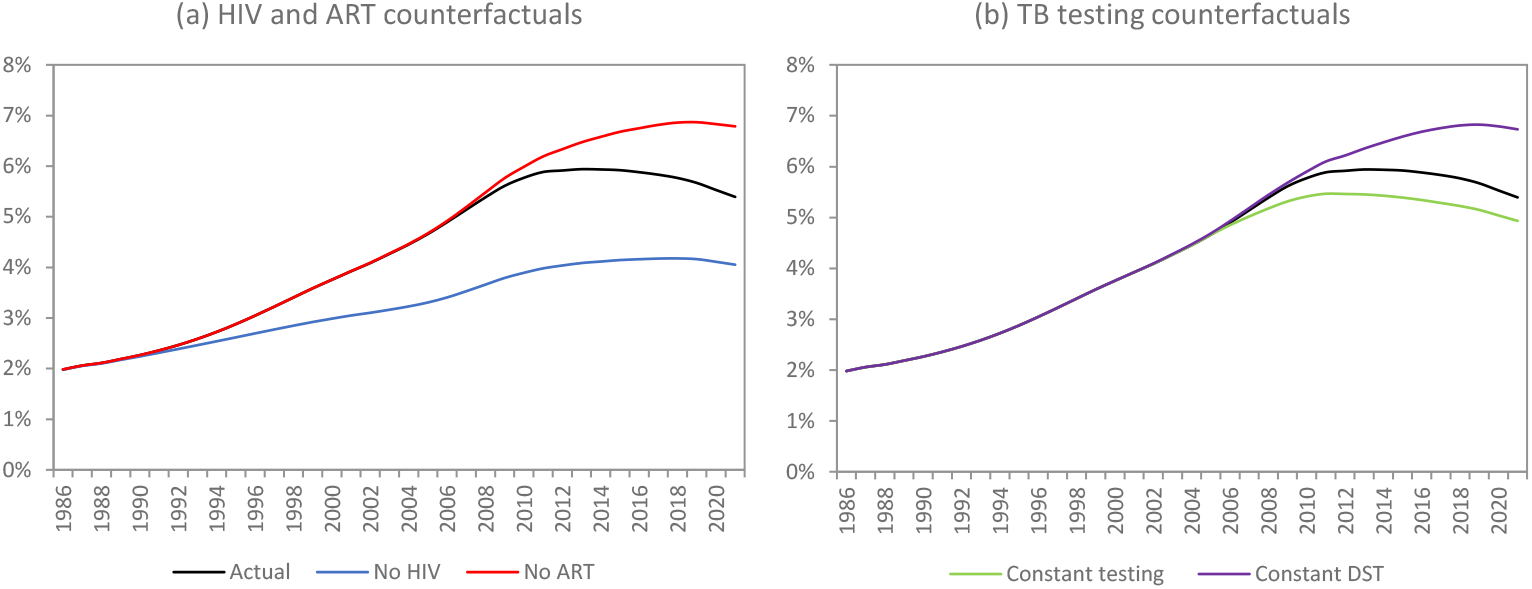
Proportion of prevalent (untreated) adult tuberculosis that is rifampicin resistant ART = antiretroviral treatment, DST = drug sensitivity testing.

Figure 4b shows that the increase in DST after 2008 has led to slight reductions in the prevalence of rifampicin resistance (by 2019 expanded DST reduced the prevalence of RR-TB by 17%). However, if general screening rates had not been increased after 2004, a *lower* prevalence of rifampicin resistance would have been expected, which is because higher rates of treatment initiation (in the absence of any change in DST) imply higher risks of acquired drug resistance.

Figure 5 shows our model estimates of the proportion of new rifampicin resistance that is acquired (while on treatment) rather than transmitted. Our model estimates that this proportion has declined from 24% (95% CI: 15-34%) in 2005 to 7% (95% CI: 5-13%) in 2020. Much of this decline is attributable to the impact of ART; in the ‘no ART’ counterfactual, the acquired fraction would have reduced only to 15% (95% CI: 10-23%) by 2020.

**Figure 5.**
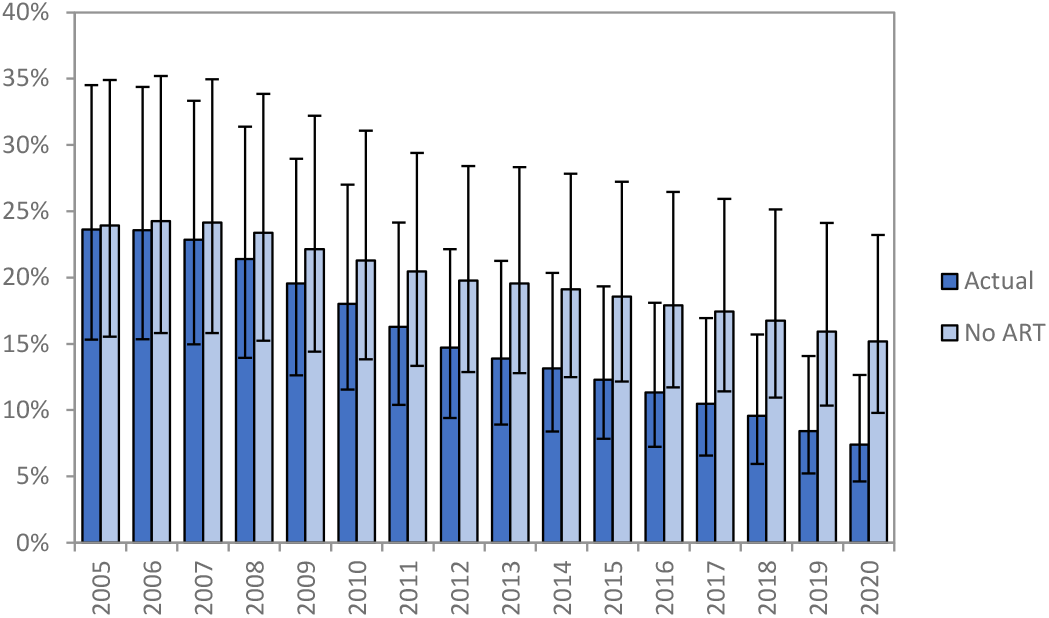
Proportion of new rifampicin-resistant cases due to acquired (not transmitted) drug resistance Error bars represent 95% confidence intervals.

## Discussion

Our results suggest that although there was a steady rise between the 1980s and 2013 in the proportion of TB that is RR-TB, this proportion has declined steadily since 2013, the time of South Africa’s last national drug-resistant TB prevalence survey [5]. This decline was partly due to the effect of ART (reversing the effect of HIV on acquired drug resistance), and partly due to increased DST in the period after 2008, with each of these factors contributing to a 17% reduction in RR-TB prevalence in 2019. Although the significance of increased DST coverage is recognized in global statistics [6], the significant contribution of ART in countries like South Africa, which have high HIV prevalence, has not previously been appreciated.

Consistent with other modelling studies [8, 36], our model suggests that acquired drug resistance is accounting for a decreasing fraction of new drug resistance over time, with transmitted resistance accounting for the vast majority of drug resistance. Levels of acquired resistance in South Africa were initially relatively high, because of the effect of HIV on the risk of acquired resistance, but the introduction of ART has contributed to a reduction in this proportion of rifampicin-resistant TB that is acquired. In settings where transmitted drug resistance accounts for most incident RR-TB, it is relatively more important to consider interventions to diagnose and appropriately treat drug-resistant TB [36], through interventions such as active case finding and improved follow-up of people diagnosed with RR-TB.

A strength of our model is that it is integrated into a detailed HIV model [37], making it well suited to addressing the primary research question. Although there have been a number of other models of RR-TB developed previously [7, 8, 35, 36, 38, 39], to our knowledge none has reflected the effect of HIV and ART on the rate of acquiring rifampicin resistance. Another strength of our model is that it distinguishes first-line and second-line TB treatment, which is important for the purpose of representing heterogeneities in outcomes and costs (a higher risk of adverse outcomes on first-line treatment [1] but higher costs of second-line treatment). With a few exceptions [39], most other models do not consider this distinction.

A limitation of our model is that we only model resistance in active TB and not in latent TB. This is potentially important when considering the provision of TPT to people with latent TB, as TPT efficacy is likely to be lower in people with latent resistant infections, but since the reduction in TPT efficacy due to resistance has not been robustly estimated, we do not attempt to model this interaction. We also implicitly assume that all transmitted RR-TB is due to ‘fast progression TB’ (developing on average one year after MTB infection) rather than reactivation. Although this assumption is not realistic, recent research suggests that reactivation accounts for only a small fraction of incident TB in high-burden settings [40], consistent with molecular evidence from South Africa [41, 42] and Malawi [43].

Our model suggests that there is likely to be substantial double-counting of RR-TB results in the NICD data, which could be due to multiple tests being performed in the same individual and test results not being adequately de-duplicated. However, our model does not always fit the calibration data well, and this finding should be treated with caution. There is a tension between fitting the survey data and the two routine data sources on rifampicin-resistant cases. For example, if we were to reduce the model estimates of rifampicin resistance prevalence to be more consistent with the survey data in 2012-14 (Figure 2), this would imply an even greater overcount in the routine NICD data. The NICD data suggest a sharp peak in RR-TB diagnosis in 2012, while the second-line treatment data suggest a later peak (in 2014-15), with our model being more consistent with the former. The advantage of the multi-evidence synthesis approach applied here is that it avoids excessive reliance on a single data source, which can lead to biased estimates.

Our model considers only rifampicin resistance. Some models simulate multiple classes of drug resistance [7, 8, 38], and these are particularly important when considering extensively drug-resistant TB (XDR-TB). Our model also considers only adult TB. The model also does not incorporate recent RR-TB programme data, and results have therefore been presented only up to 2020. Because of this, we have not considered here the impact of newer RR-TB drug regimens [44], which are both shorter and more efficacious. Further analysis of the costs of screening for and treating RR-TB are ongoing [45], and will be important in informing future evaluations of the most cost-effective strategies to limit RR-TB.

Despite these limitations, our study is important in demonstrating the importance of ART in reducing the incidence of RR-TB. In the face of recent threats to HIV and TB programme funding [46], it is critical that ART programmes are not compromised and that DST is sustained.

## Data Availability

All data produced in the present study are available upon reasonable request to the authors. Most of the data used in the model calibration are from publicly available sources cited in the paper.

## Appendix

### Calculation of proportion of TB patients with previous TB treatment

For the purpose of calculating the number of TB patients who receive DST in the period before GeneXpert rollout, it is necessary to first estimate the proportion who have a history of prior TB treatment. Suppose that in year *t*, the number of new drug-sensitive and new drug-resistant TB cases are *DS*_*h*_(*t*) and *DR*_*h*_(*t*) respectively, with treatment history *h* (*h* is 0 if never treated before, or 1 if treated previously). If *DT*_*h*_(*t*) is the total number of new TB cases in year *t*, with treatment history *h*, then DR_*h*_(*t*) is calculated using the previous formula for determining the proportion of incident cases that are drug-resistant, i.e.

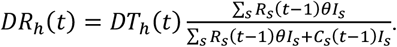

Further suppose that the number of TB patients who experience treatment failure in year *t*, with drug resistance, is *FR*(*t*), and the number who experience treatment failure without drug resistance is *FS*(*t*) (in both cases we include in the definition of treatment failure only those who stop treatment, i.e. excluding those who continue a longer course of treatment). Then we approximate the proportion of treated drug-resistant TB cases who are previously treated as

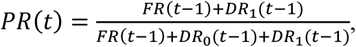

and similarly the proportion of newly treated drug-susceptible TB cases who are previously treated is

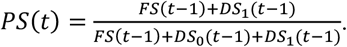

Again, we are using a time lag of one year, which is to take account of the delay between when active TB develops and when treatment is initiated. This is a crude approximation, because in most of our model experiments, the average prevalence-to-incidence ratio is substantially greater than 1 (suggesting a delay of more than one year). The implicit assumption being made here is that people who newly acquire TB and those who experience TB treatment failure seek treatment (or re-treatment) at the same rate.

### Likelihood definition

#### Survey data

Our model is calibrated to the results from two national drug resistance surveys in TB patients [5, 12]. Both surveys were conducted in patients presenting with TB symptoms (i.e. prior to initiating treatment). The first survey, conducted in 2001-02, found an overall rifampicin resistance prevalence of 3.4% (95% CI: 2.5-4.3%) in patients with microbiologically confirmed TB. The second survey, conducted in 2012-2014, found the prevalence of rifampicin-resistant TB was 4.6% (95% CI: 3.5-5.7%) overall.

As noted in the main text, a limitation of both surveys is that they relied on phenotypic detection methods that missed “borderline resistant mutations” that were not considered defining of resistance at the time, but which are now included in the WHO definition of rifampicin resistance (Shaheed Omar, personal communication). The exact proportion of rifampicin resistant cases that would have been missed is unknown, as it is highly variable across settings, but in a global survey the average proportion was 12% (Shaheed Omar, personal communication). The measured rifampicin resistance prevalence levels (3.4% and 4.6%) has therefore been adjusted to account for an assumed 88% sensitivity (to 3.9% and 5.2% respectively).

We compare the model estimate of the proportion of untreated adults with TB who have rifampicin resistance, in mid-2001 and mid-2013, to the adjusted survey estimates of 3.9% and 5.2% respectively. For the sake of defining the likelihood for the survey data, we assume that the difference between the modelled rifampicin-resistant proportion and the observed proportion, on a logit scale, is normally distributed with zero mean. The standard deviation of the normal distribution is calculated from the 95% confidence intervals quoted previously.

#### NICD datanumbers of rifampicin-resistant diagnoses

Our model is also calibrated to recorded numbers of patients who are diagnosed with rifampicin-resistant TB (which are based both on specimens collected prior to treatment initiation and specimens collected in patients who are not responding to treatment). These data are reported in a number of sources. The most recent data for 2020 and 2021 were obtained directly from the National Health Laboratory Service (NHLS). Data for the period 2010-2019 are obtained from the World Health Organization (WHO) reporting system [29], which draws on earlier NICD reporting. Although data are also reported before 2010 [26], these are inconsistent with the more recent WHO data in the period after 2010, and are therefore not included in the model calibration.

The NHLS estimates could be over-estimates or under-estimates of the true numbers of diagnoses. The reporting system is not based on a unique patient ID, but rather on a probabilistic matching algorithm for linking different laboratory results to the same patient. There is thus potential for under-matching (implying over-estimated numbers of diagnoses) or over-matching. There is also potential for diagnostic error: in 2020 and 2021, only about half of the rifampicin-resistant cases identified by Xpert were confirmed by culture or LPA. Specificity of Xpert and Xpert Ultra, in the detection of rifampicin resistance, is estimated to be around 99% [47]. It is also possible that the sensitivity of Xpert in detecting rifampicin resistance may be poor in routine settings – Zurcher *et al* [1], for example, estimate a sensitivity of only 80%. However, we would expect any bias due to mis-matching or diagnostic inaccuracy to be relatively stable over the 2010-2021 period.

We therefore specify the likelihood for the NHLS data on the assumption that the bias, on the log scale, is constant over the 2010-2020 period. If *Y*(*t*) is the modelled number of rifampicin-resistant diagnoses in year *t*, and *L*(*t*) is the corresponding recorded number, then we specify the likelihood on the assumption that 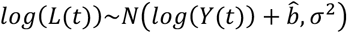, where

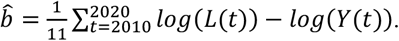

The standard deviation *σ* is set to 0.1 on the log scale, which is equivalent to a standard deviation that is approximately 10% of the true number of diagnosed drug-resistant cases on the normal scale. It is worth noting that the model projection years run from mid-year to mid-year, whereas the NHLS data relate to calendar years; we therefore average the NHLS data across successive years for the purpose of calculating the likelihood. (For example, we compare the model estimate for the 2010-2011 projection year to the average of the NHLS data in 2010 and 2011.)

#### EDRweb data: numbers of patients starting second-line treatment

Finally, our model is calibrated to recorded numbers of patients starting second-line treatment. Data for the period 2007-2020 period are obtained from the WHO reporting system [26, 29], based on EDRweb, the national electronic reporting system for drug-resistant TB. These reported numbers are likely to be under-estimates, as the reporting challenges facing EDRweb are similar to those for the Electronic TB Register (ETR), the register for drug-susceptible TB. Many patients starting TB treatment, especially in hospital settings, do not get recorded on ETR [48-50], and cases treated in the private sector (which may account for as much as 15% of treated TB [51]) are also not included in ETR.

In specifying the likelihood for the EDRweb data, we assume that the bias due to this undercount is (proportionally) constant over time. Similar to the previous specification, if *X*(*t*) is the modelled number of second-line treatment initiations in year *t*, and *E*(*t*) is the corresponding recorded number, then we specify the likelihood on the assumption that *log*(*E*(*t*))~*N*(*log*(*X*(*t*)) + ĝ, *σ*^2^), where

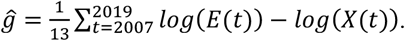

The standard deviation *σ* is again set to 0.1 on the log scale. As before, we average *E*(*t*) across successive calendar years in order to align these with the Thembisa projection years.

